# Implicit Competition and Serotype Replacement: Insights from Mathematical Modeling of Multi-colonization and Vaccination

**DOI:** 10.1101/2025.07.15.25331574

**Authors:** Tufail Malik, Oluwaseun Sharomi, Elamin H Elbasha

## Abstract

In this research, we investigate the dynamics of interactions among multiple serotypes of an infectious agent in presence of vaccination and reveal insights into the phenomenon of serotype replacement. Traditionally, serotype replacement following vaccination has been interpreted primarily as a consequence of direct competition among different serotypes. However, our study demonstrates that these observed shifts can also stem from the modeling constraints on multi-colonization. We provide evidence that, even in the absence of explicit competitive interactions between serotypes, modeling assumptions on the number of serotypes allowed to co-colonize can inadvertently foster an implicit competition that exhibits serotype replacement ensuing vaccination. We further support this argument by examining pneumococcal multi-colonization dynamics as a case study. This finding challenges the conventional understanding of serotype dynamics, suggesting that model-projected occurrence of replacement following vaccination is not solely an epidemiological event driven by serotype competition but can also be an artifact arising from the limitations of the modeling framework. Understanding these dynamics is essential for developing more accurate predictive models and devising effective public health interventions.

## 1 Introduction

At the host level, individuals can be co-colonized with multiple serotypes of bacterial or viral species simultaneously (i.e., they can coinfect a host). SARS-Cov-2 [3, 4, 15] and Pneumococcus [10, 14] are two of the many examples of co-circulating and sometimes coinfecting agents. The two phenomena of multiple serotypes existing in the population and within a host are distinguished in Martcheva et al. (2006) [13], and are termed as coexistence and coinfection, respectively. The model presented in [13] has no coexistence equilibria without coinfection. On the other hand, coinfection may support multiple coexistence equilibria. Coexistence, when feasible, is only possible for small values of reproduction numbers, otherwise one disease dominates.

Lipsitch (1997) [11] establishes conditions for coexistence and competitive exclusion of serotypes under mass vaccination and shows that with a monovalent vaccine, the increase in the prevalence of a nonvaccine type (NVT) in a 2-serotype model will not exceed the amount of decline of the vaccine-type (VT), which is not necessarily the case in a 3-serotype model. Elbasha *et al*. (2005) [7] present a two HPV type vaccination model and derive conditions for a coexistence equilibrium in presence of a full-protection monovalent vaccine.

The current paper investigates the conditions that facilitate coexistence of multiple serotypes and the drivers of serotype replacement following vaccination within a standard compartmental model. It then uses multi-colonization (MC) in pneumococcal carriage as a case study to demonstrate that serotype replacement observed in modeling studies may be an artifact of modeling assumptions of MC, and not necessarily due to the direct serotype competition. *Streptococcus pneumoniae* (SP) is a major cause of morbidity and mortality in children and adults worldwide [16]. It is known that asymptomatic nasopharyngeal carriage plays an essential role in the transmission of *Streptococcus pneumoniae* [1]. There are over 100 serotypes of *Streptococcus pneumoniae*, with varying degree of propensity to causing pneumococcal disease in humans. Many serotypes circulate in a population simultaneously (i.e., establish coexistence equilibria). Furthermore, data on unvaccinated children colonized with SP serotypes [2] showed a 10-fold reduction in the probability of acquisition of another SP serotype in the colonized individuals. This is an evidence of decreased susceptibility to acquiring a specific serotype when an individual is already colonized with a different serotype, compared to someone who is completely susceptible to both serotypes (i.e., there is serotype competition).

The widespread use of pneumococcal conjugate vaccines (PCVs) since 2000 has significantly impacted the ecology of SP and the resulting disease in vaccinated and unvaccinated individuals across the age span [17, 18]. This included both herd protection effect and serotype replacement.

## 2 Multiple-serotype vaccination model

Antigenically different serotypes can co-colonize a host. This motivates the construction of a multi-serotype model where a host can be infected with and carry multiple serotypes at the same time.

We divide the total population at time *t, N* (*t*), into distinct compartments based on colonization with single or multiple serotypes from the set *X* = *{*1, 2, …, *n}* and vaccination status. Each serotype will be denoted by *ST*_*i*_, *i ∈ X*. The unvaccinated population consists of susceptible individuals, denoted as *S*(*t*) and individuals infected with a single *ST*_*i*_, denoted as *I*_*i*_(*t*) and those colonized with two or more serotypes in the subset **h** of *X*, denoted as *I*_**h**_(*t*), for **h P**(*X*) \ *{x}, x ∈ X*, where **P** denotes the power set. The corresponding compartments of vaccinated individuals, respectively, are denoted as *V* (*t*), *W*_*i*_(*t*) (where *i ∈ X*) and *W*_**h**_(*t*) (where **h P**(*X*) \ *{x}, x ∈ X*). Infection with and recovery from multiple serotypes are assumed to be sequential. That is, when a co-colonized host comes into contact with a susceptible host, only one of the serotypes can be transmitted. Also, a co-colonized host clears one serotype at a time. The model additionally assumes that serotypes directly compete for the same resources, which can reduce the ability of one serotype to invade a niche already colonized by another serotype.

The transmission dynamics of colonizing pneumococcal bacteria in a homogenously mixing population are commonly modeled using a standard susceptible-infected-susceptible (SIS) formulation [8, 9, 11]. Thus, no immunity or cross-immunity following colonization is assumed. By adopting this compartmental structure, we aim to examine the dynamics of pneumococcal transmission and coexistence within a population in presence of serotype competition, considering the specific context of two serotypes. The resulting model, depicted by Figure 1, is given by the following deterministic system of nonlinear ODEs (where a dot represents differentiation with respect to time *t*):

**Figure 1:**
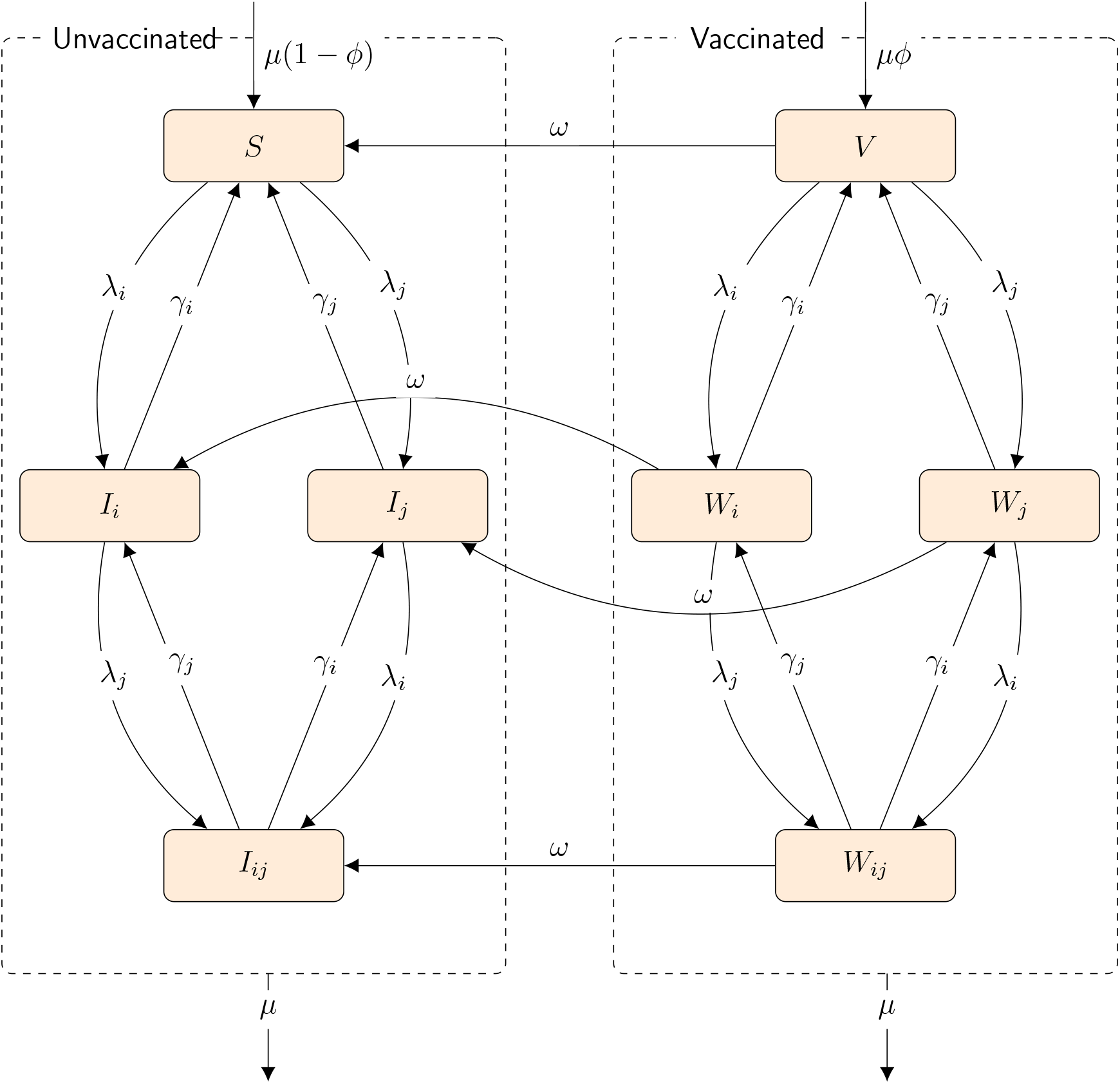
Flowchart of the model representing the transmission dynamics of pneumococcal bacteria in a homogeneously mixing population, focusing on two serotypes, *i* and *j*. The population is divided into compartments based on colonization status (susceptible, infected with a single serotype, or co-colonized) and vaccination status. Susceptible Individuals: Denoted as *S* for the unvaccinated and *V* for the vaccinated. Unvaccinated infected Individuals: *I*_*i*_ and *I*_*j*_ represent individuals infected with serotypes *i* and *j*, respectively, while *I*_*ij*_ denotes those co-colonized with both serotypes. Vaccinated Infected: *W*_*i*_ and *W*_*j*_ represent vaccinated individuals infected with serotypes *i* and *j*, respectively, and *W*_*ij*_ for those co-colonized.

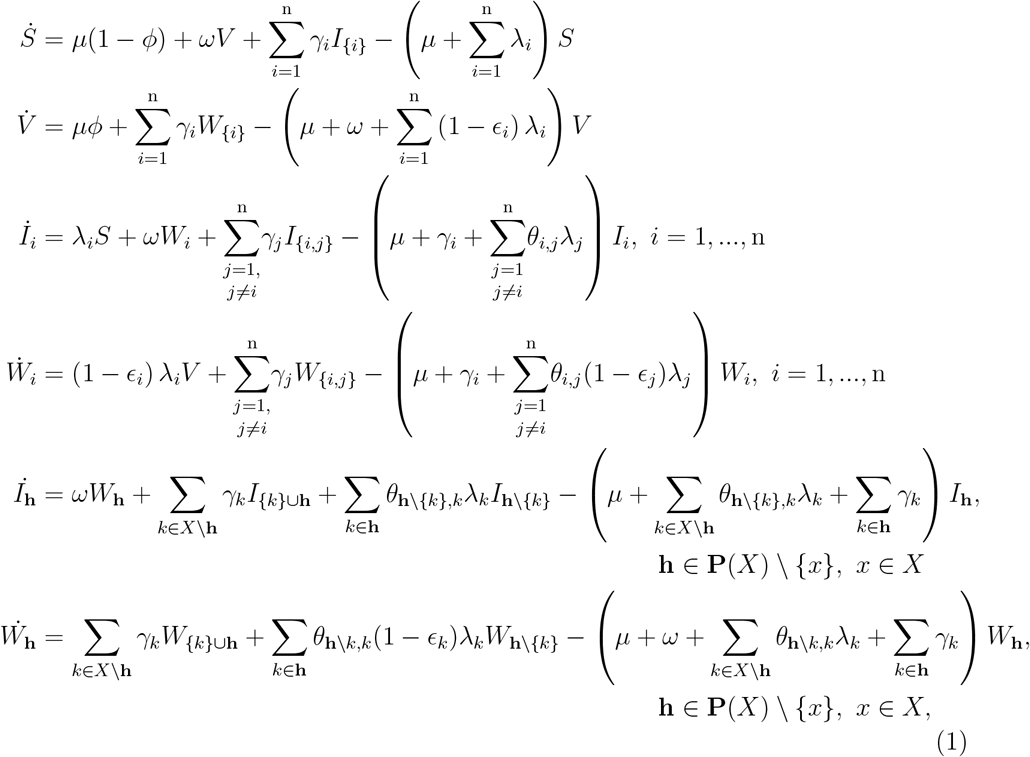

where 0 *≤ ϵ*_*i*_ *≤* 1 is the degree of protection conferred by the vaccine against infection with serotype *i*. It is assumed that a fixed proportion, *ϕ*, of newborn is vaccinated for an average protection period of 1*/ω*.

The forces of infection are given by,

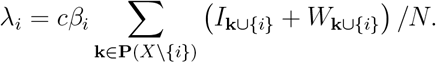

The force of infection for serotype *i* quantifies the rate at which individuals become infected with serotype *i*. The parameter *µ* corresponds to the per-capita birth and natural death rate, accounting for the rate at which individuals enter and exit the population due to birth and death. The parameter *γ*_*i*_ denotes the rate of recovery from infection with serotype *i*, indicating the speed at which individuals clear the infection and return to a susceptible state. In this context, *β*_*i*_ represents the probability of transmission of serotype *i* per contact, and *c* represents the average number of contacts an individual has during a specific time period. The metabolic-type competition is captured by the parameter *θ*_*i,j*_ (or *θ*_**h**,*j*_), which represents the reduction in fitness or acquisition efficiency of serotype *j* if an individual is already colonized with the another serotype *i* (or a group of serotypes **h, h** *∈* **P**(*X*) *\ {j}*). This parameter satisfies the condition 0 *< θ*_*i,j*_ *<* 1 (similarly, 0 *< θ*_**h**,*j*_ *<* 1). When *θ*_*i,j*_ = 1 (or, similarly, *θ*_**h**,*j*_ = 1), there is no competition, and when *θ*_*i,j*_ = 1 (or, similarly, *θ*_**h**,*j*_ = 0), competitive exclusion of serotype *j* occurs.

It is possible to compute explicit expressions for the prevalence of serotypes within models containing two or three serotypes. Consequently, we will first examine a model with two serotypes, followed by an analysis involving three serotypes, to investigate how the prevalence of coexisting serotypes affects prevalence of individual serotypes.

## 3 Two-serotype model with a limited duration, fully protective vaccine

Model (1) involving two serotypes, depicted in Figure 1, can be explicitly written as

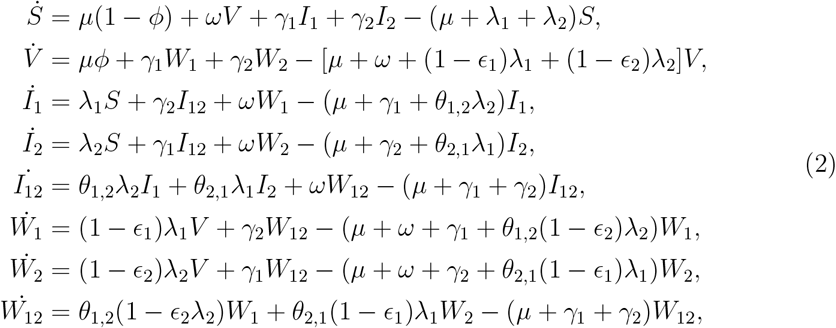

The forces of infection are given by

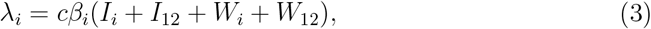

Here, the model (2) with a monovalent (*ST*_1_-specific) vaccine with 100% efficacy (i.e., *ϵ*_1_ = 1, *ϵ*_2_ = 0) is analyzed. Results for the case without vaccination can be obtained by setting the vaccination rate to zero (i.e., *ϕ* = 0). The model (2) has four different feasible equilibria, *E*_*j*_, *j* = 0, 1, 2, 3. The population size *N* is assumed to remain constant and normalized to 1, with births balancing the deaths in the population.

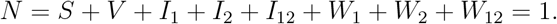

This assumption allows us to focus on the dynamics of serotype MC and transmission without considering population growth or decline.

Since the total population is constant at equilibrium, it is sufficient to consider the dynamics of the flow generated by the model (2) in the positively invariant region

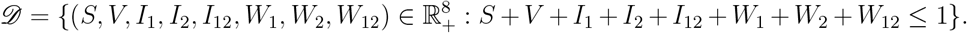

It can be shown that unique solutions exist in *𝒟* for all positive time. Therefore, the model is epidemiologically and mathematically well posed [9].

The model (2) has four different feasible equilibria: the disease-free equilibrium, two boundary endemic equilibria (with one of the two serotypes present) and coexistence endemic equilibrium with both serotypes present. The reproduction numbers for disease-free equilibrium to be invaded by *ST*_*i*_ are given, respectively, by [5, 6].

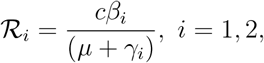

The basic reproduction number for the model (2) is given by *R*_0_ = max(*ℛ*_1_, *ℛ*_2_), where *ℛ*_1_, *ℛ*_2_ represent the average number of secondary cases of carriage of *ST*_1_ and *ST*_2_, respectively, generated by a typical individual carrying the corresponding serotypes during their carriage period, when introduced into a completely noncolonized susceptible population in the absence of a pneumococcal vaccine [9].

It can also be shown that, compared with the case of no competition (i.e., *θ*_1,2_ = *θ*_2,1_ = 1), competition always results in lower equilibrium prevalence. With prevalence defined as *p*_*i*_ = *I*_*i*_ + *I*_12_, model (2) can be solved for endemic equilibrium as

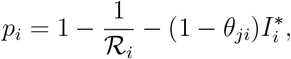

*i* ≠ *j, i*, 1, 2; *j* = 1, 2. Clearly, when *θ*_*ji*_ *<* 1, *p*_*i*_ is lower. This does not necessarily imply that an increase in competition always lead to lower prevalence of both serotypes. To illustrate consider the polar case where *θ*_12_ = 0, but 0 *< θ*_21_ *<* 1. In this case, equilibrium prevalence is given by

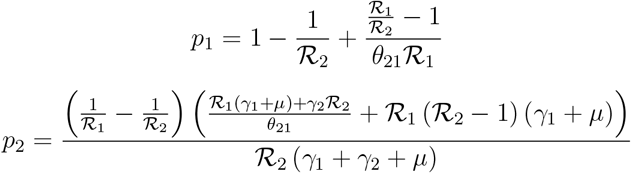

Thus, because *ℛ*_2_ *> ℛ*_1_, *p*_1_ goes down but *p*_2_ goes up with more competition (i.e., lower values of *θ*_21_). In this case, starting from an equilibrium with a given level of competition, more competition implies less prevalence of serotype 1 but more prevalence of serotype 2.

The model (2) with a monovalent (*ST*_1_-specific) vaccine with 100% efficacy (i.e., *ϵ*_1_ = 1, *ϵ*_2_ = 0) has four different feasible equilibria, *E*_*j*_, *j* = 0, 1, 2, 3.

### 3.1 Disease-free equilibrium

The unique disease-free equilibrium is given by

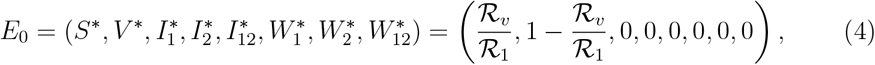

where 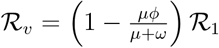 is the effective reproduction number of *ST*_1_. The Jacobian matrix evaluated at the disease-free equilibrium (4) (see Appendix) has eigenvalues:

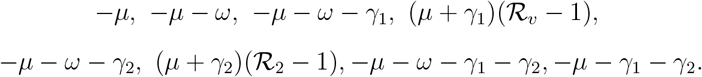

Thus, we have the following result.

#### Theorem 3.1

*E*_0_ *is locally-asymptotically stable (i*.*e*., *all eigenvalues are negative) if R*_0_ = max *{ℛ*_*v*_, *ℛ*_2_*} <* 1. *Otherwise, E*_0_ *is unstable*.

### 3.2 Boundary equilibria

The equilibrium where only *ST*_1_ is endemic is given by

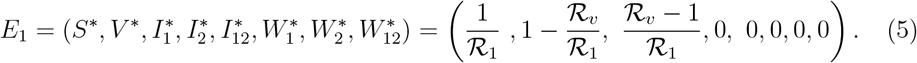

*E*_1_ exists only if *ℛ*_*v*_ *>* 1. The Jacobian matrix evaluated at the disease equilibrium (5) (shown in the Appendix) has eigenvalues:

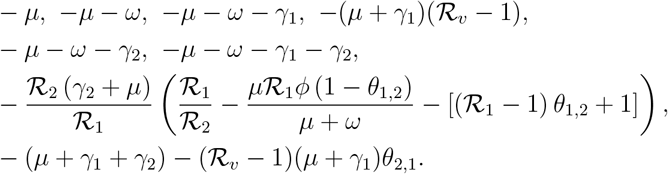

Thus, *E*_1_, when it exists, is locally stable (i.e., all eigenvalues are negative) provided

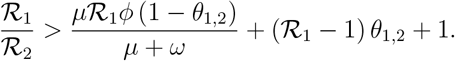

The equilibrium where only *ST*_2_ is endemic is given by

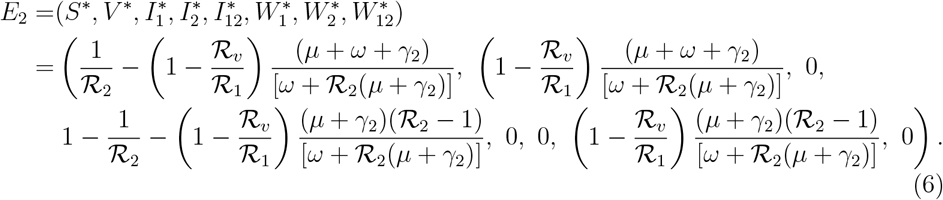

*E*_2_ exists only if *ℛ*_2_ *>* 1.

The Jacobian matrix evaluated at the disease equilibrium (6) (also shown in the Appendix) has eigenvalues:

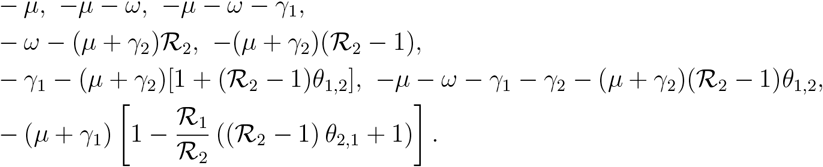

*E*_2_, when it exists, is locally stable (i.e., all eigenvalues are negative) provided

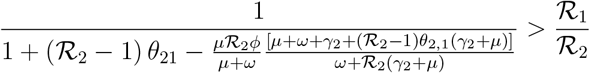

Otherwise, *E*_2_ is unstable.

The theoretical results established in this section are summarized below.

#### Theorem 3.2

*The model (2) has a unique ST*_1_*-only (ST*_2_*-only) boundary equilibrium only if ℛ*_*v*_ *>* 1 *(ℛ*_2_ *>* 1*), and no boundary equilibrium otherwise*.

#### Theorem 3.3

*The boundary equilibrium E*_1_ *is locally-asymptotically stable if*

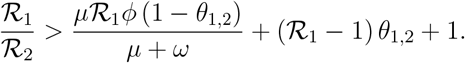

*Otherwise, E*_1_ *is unstable*.

#### Theorem 3.4

*The boundary equilibrium E*_2_ *is locally-asymptotically stable if*

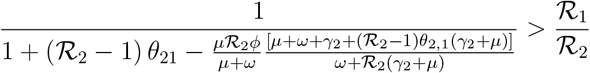

*Otherwise, E*_2_ *is unstable*.

### 3.3 Coexistence endemic equilibrium

The coexistence equilibrium where both serotypes are endemic occurs when both single-type endemic equilibria (*E*_1_ and *E*_2_) exist and are unstable.

#### Theorem 3.5

*The model (2) has a locally-asymptotically stable coexistence equilibrium whenever* min*{ℛ*_*v*_, *ℛ*_2_*} >* 1, *and*

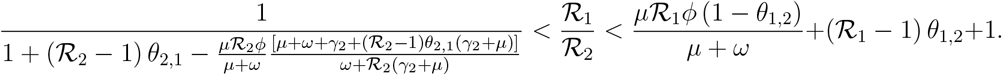

By assuming independence in transmission and clearance, and in the presence of MC, the interaction between the two serotypes is determined entirely by the competition parameters *θ*_1,2_ and *θ*_2,1_. In the absence of competition (i.e., *θ*_1,2_ = 1 and *θ*_2,1_ = 1), prevalence in the coexistence endemic equilibrium with a monovalent, perfect vaccine has a simple structure:

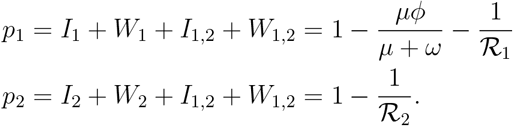

Thus, we conclude the following:

#### Theorem 3.6

*In the absence of competition (i*.*e*., *θ*_1,2_ = *θ*_2,1_ = 1*) among serotypes of the model (2), the prevalence of each serotype at the coexistence endemic equilibrium, when a monovalent and perfect vaccine (i*.*e*., *ε*_1_ = 1, *ε*_2_ = 0*) is administered, is determined solely by the intrinsic parameters of each individual serotype*.

In particular, there is no interference from parameters related to *ST*_1_ on the equilibrium prevalence level of the non-vaccine type *ST*_2_ after the decline in *ST*_1_ following vaccination (note that higher vaccination coverage rate (VCR) or longer duration of vaccine protection results in lower levels of equilibrium *ST*_1_). In other words, there is no serotype replacement.

## 4 Three-serotype vaccination model without direct competition

We have established that in the absence of parameter-based interaction between serotypes (independence of transmission and clearance, and metabolic-type competition) in a two-serotype model with MC, prevalence of each serotype at endemic equilibria is not influenced by the other serotype. In other words, serotype replacement is ruled out. Indeed, replacement is possible when serotypes compete for resources (i.e., if metabolic-type competition exists).

By considering a third serotype, the implications of using different assumptions regarding the ability to be colonized by multiple serotypes can be examined. Therefore, we will focus on the model described in equation (1) that incorporates three serotypes (*ST*_1_, *ST*_2_ and *ST*_3_) which we will subsequently refer to as the 3-ST model. With a monovalent, perfect vaccine (i.e., *ε*_1_ = 1, *ε*_2_ = 0, *ε*_3_ = 0, where an index *i* represents ST*i*) in the absence of serotype competition (i.e., *θ*_*ij*_ = *θ*_*ijk*_ = 1, *i ≠j, j≠k*; *i, j, k* = 1, 2, 3) the 3-ST model has several equilibria, including the equilibrium where the three serotypes coexist. We will analyze the model with and without triple colonization.

### 4.1 Serotype prevalence with level 3 MC

Here we assume that individuals can get colonized with all three circulating serotypes. Adding up the equations that include serotype 2 infectious compartments, defining serotype 2’s prevalence by *p*_2_, and utilizing the total population equation, it can be shown that

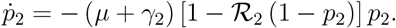

In equilibrium,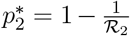. Clearly, the dynamic behavior of *P*_2_ is entirely determined by *µ* and the parameters of *ST*_2_, *γ*_2_ and *ℛ*_2_. There is no ST replacement in this case. Similarly, the prevalence of *ST*_3_ is given by

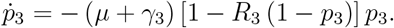

By adding up the equations that include *ST*_1_ infectious compartments, defining *ST*_1_’s prevalence by *p*_1_, and utilizing the total population equation, it can be shown that

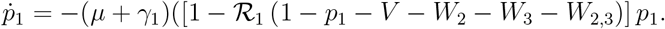

The ODE for *V* + *W* _2_ + *W*_3_ + *W*_2,3_ (noting that there are no breakthroughs) are given by 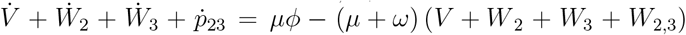. This can be solved analytically as

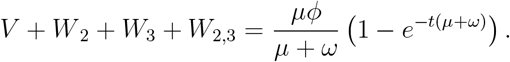

Thus,

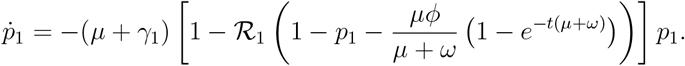

Clearly, the dynamic behavior of *p*_1_ is entirely determined by vaccine properties *ϕ* and *ω, µ* and the parameters of *ST*_1_, *γ*_1_ and *ℛ*_1_.

The results of this section are summarized in the following theorem.

#### Theorem 4.1

*Consider the model (1) with three serotypes and level 3 MC. With a monovalent, perfect vaccine (ε*_1_ = 1, *ε*_2_ = 0, *ε*_3_ = 0, *where an index i represents ST*_*i*_*) in the absence of serotype competition (θ*_*ij*_ = *θ*_*ijk*_ = 1, *i ≠ j, j≠ k*; *i, j, k* = 1, 2, 3*) the prevalence of each serotype at the coexistence equilibrium (where all three serotypes coexist) is determined solely by the intrinsic parameters of each individual serotype*.

### 4.2 Serotype prevalence with level 2 MC

This case assumes that individuals can be co-colonized with only two serotypes. That is, in the 3-serotype model, we set *I*_123_ = *W*_123_ = 0.

In the endemic equilibrium the prevalence of serotype 2 can be computed as

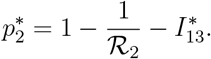

Similarly, the endemic prevalence of serotype 3 can be computed as

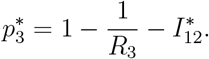

Finally, the endemic prevalence of serotype 1 is

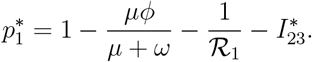

Compared to the case when triple colonization is possible, the endemic equilibrium value of 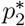 now also depends on the prevalence of colonization of serotype 1 and 3. It is also lower when triple colonization is not allowed.

These results are captured in the following.

#### Theorem 4.2

*Consider the model (1) with three serotypes and level 2 MC. With a monovalent, perfect vaccine (ε*_1_ = 1, *ε*_2_ = 0, *ε*_3_ = 0, *where i represents ST*_*i*_*) in the absence of serotype competition (θ*_*ij*_ = *θ*_*ijk*_ = 1, *i* ≠ *j, j* ≠ *k*; *i, j, k* = 1, 2, 3*) the prevalence of each serotype at the coexistence equilibrium (where all three serotypes coexist) depends on prevalence of other serotypes*.

We conclude that with triple colonization, there is no ST-replacement in the absence of explicit serotype competition. However, in the absence of triple colonization, prevalence of serotypes at equilibrium is interfered by prevalence of other serotypes. This suggests that in a 3-ST model with only level 2 MC, serotype replacement can also occur even in the absence of direct serotype competition.

To further investigate possible serotype replacement following vaccination in models with higher number of serotypes, we numerically compare prevalence of serotypes with and without vaccination.

## 5 Serotype replacement without direct competition: a case study of pneumococcal vaccines

In this section, we utilize the dynamics of pneumococcal carriage to numerically demonstrate that the model may project prevalence of non-vaccine serotypes to increase as the prevalence of vaccine-targeted serotypes decreases due to vaccination, even in the absence of direct competition between serotypes (i.e., with *θ*_*i,j*_ = 1 for all *i, j*). This phenomenon arises from the model’s limitations on the number of serotypes that can cocolonize at the same time. Pneumococcal conjugate vaccines (PCVs) in children have been progressively expanded from previous formulations by incorporating additional serotypes. The sequence of PCV introductions includes the 7-valent vaccine PCV7, followed by the 13-valent PCV13, the 15-valent PCV15, and finally, the 20-valent PCV20. A dynamical transmission model of pneumococcal carriage was introduced by Malik *et al*. [12] that accounts for six distinct age groups, eleven serotype classes (STCs, grouping serotypes according to vaccines), and the previously mentioned PCVs, along with V116 and PPSV23. Permitting MC with all eleven STCs in that model would be mathematically impractical. This would introduce significant complexity due to nonlinearities and considerably higher number of model compartments, leading to extensive demands on computational resources and time, rendering any analytical efforts nearly unfeasible.

In contrast, the simpler framework of the model (1) (such as no age stratification and no multiple serotype transmission) facilitates a comparative analysis of the effects of restricting MC on serotype replacement. We designate the 11 STCs as 11 distinct serotypes, denoted as *ST*_1_, *ST*_2_, …, *ST*_11_. In this notation, PCV7 targets *ST*_1_ (representing all serotypes targeted by PCV7), PCV13 targets *ST*_1_ through *ST*_2_ (where *ST*_2_ represents additional serotypes targeted by PCV13), and so forth. We set the initial values of model compartments as *S*(0) = 0.6, *V* (0) = *W*_*i*_(0) = 0, *I*_*i*_(0) = 0.4*/*5 for all individual serotypes *i*, and *W*_*j*_(0) = *I*_*j*_(0) = 0 for any co-colonized compartments with serotypes group *j*. Vaccine efficacy of each PCV against a target serotype is listed in Table 1, with identical efficacies of all PCVs targeting a given ST.

**Table 1:**
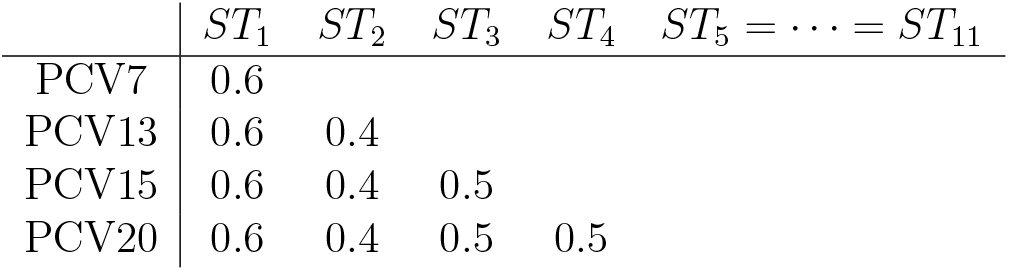
Efficacy of PCVs against target serotypes. An empty cell indicates that the serotype listed in the corresponding column is a non-vaccine type for the PCV listed in the corresponding row.

| | $ST_1$ | $ST_2$ | $ST_3$ | $ST_4$ | $ST_5 = \dots = ST_{11}$ |
| --- | --- | --- | --- | --- | --- |
| PCV7 | 0.6 |  |  |  |  |
| PCV13 | 0.6 | 0.4 |  |  |  |
| PCV15 | 0.6 | 0.4 | 0.5 |  |  |
| PCV20 | 0.6 | 0.4 | 0.5 | 0.5 |  |

Figure 2 illustrates the time-evolution of prevalence *p*_*i*_ of each *STi* following the implementation of PCV15 and PCV20. It is observed that vaccine-targeted serotypes demonstrate a decline: specifically, a reduction in *ST*_1_ through *ST*_3_ is noted with PCV15, and *ST*_1_ through *ST*_4_ decline with PCV20. When MC is restricted to only two serotypes (i.e., level 2 MC), a marked increase in the prevalence of non-vaccine types is evident upon the introduction of a PCV (left panel). However, when the model accommodates level 11 MC, non-vaccine types sustain their prevalence at levels comparable to those observed in the absence of vaccination for each respective PCV (right panel).

**Figure 2:**
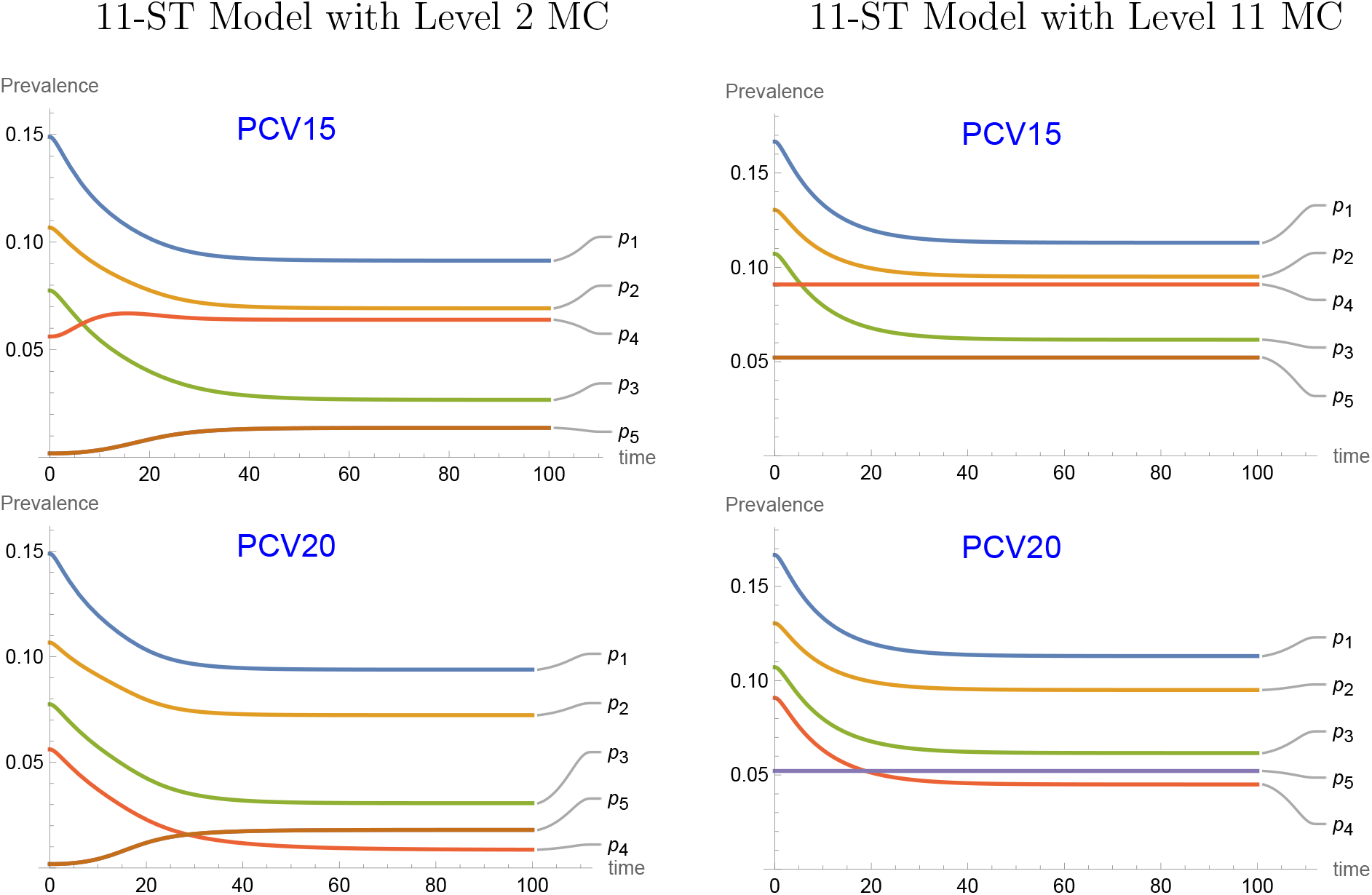
Serotype prevalence in the 11-serotype model assuming level 2 multicolonization (left column) versus level 11 multi-colonization (right column). *p*_5_, …, *p*_11_ are identical (hence, only *p*_5_ is shown). *θ*_*ij*_ = 1 for all *i, j*. Other parameter values used are: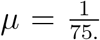, *c* = 20, *β*_1_ = 0.3139, *β*_2_ = 0.3629 *β*_3_ = 0.3795, *β*_4_ = 0.4790, *β*_5_ = *· · ·* = *β*11 = 0.5512, 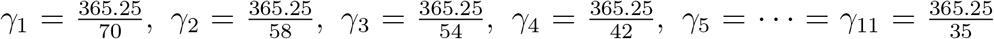 (so that, *ℛ*_1_ = 1.2, *ℛ*_2_ = 1.15, *R*_3_ = 1.12, *R*_4_ = 1.1, *R*_5_ = *· · ·* = *R*_11_ = 1.055). MC: multi-colonization.

A detailed quantitative comparison of prevalence of each serotype at equilibrium provides further insights. It is instructive to observe serotype replacement (change in prevalence of NVTs between initial and equilibrium levels). The first row of Table 2 for each level of MC displays the steady-state prevalence of each serotype in the absence of vaccination. We first consider the case where the model (1) incorporates level 2 MC. The introduction of PCV7 leads to a decrease in the prevalence of *ST*_1_. However, the other serotypes maintain their prevalence levels as observed in the no-vaccine steady state, as PCV7 does not provide protection against them. *ST*_2_ increases significantly from 0.0804 to 0.0889, representing a 26.25% rise. Additionally, the prevalence of serotypes *ST*_3_ through *ST*_5_ also increases with PCV7. Each vaccine results in a rise in the prevalence of any serotype not included in that vaccine.

**Table 2:** Steady-state serotype prevalence in an 11-serotype model with level 2, 3, 4, or 11 multi-colonization. Values in parentheses indicate the percent change in prevalence at steady state. Parameter values are consistent with those presented in Figure 2. (level *i* MC: Multi-colonization with up to *i* serotypes.)

| | $p_1$ | $p_2$ | $p_3$ | $p_4$ | $p_5 = \dots = p_{11}$ |
| --- | --- | --- | --- | --- | --- |
| level 2 MC |  |  |  |  |  |
| No Vaccine | 0.15 | 0.11 | 0.08 | 0.06 | 0.002 |
| PCV7 | 0.09(-41%) | 0.11(0.91%) | 0.08(3%) | 0.06(5%) | 0.006(243%) |
| PCV13 | 0.09(-40%) | 0.07(-38%) | 0.08(5%) | 0.06(8%) | 0.01(410%) |
| PCV15 | 0.09(-39%) | 0.07(-35%) | 0.03(-65%) | 0.06(14%) | 0.01(637%) |
| PCV20 | 0.09(-37%) | 0.07(-32%) | 0.03(-60%) | 0.01(-85%) | 0.02(866%) |
| level 3 MC |  |  |  |  |  |
| No Vaccine | 0.16 | 0.12 | 0.09 | 0.08 | 0.33 |
| PCV7 | 0.10(-35%) | 0.12(1%) | 0.10(2%) | 0.08(2%) | 0.04(7%) |
| PCV13 | 0.10(-35%) | 0.08(-30%) | 0.10(3%) | 0.08(4%) | 0.04(11%) |
| PCV15 | 0.10(-34%) | 0.09(-28%) | 0.05(-47%) | 0.08(6%) | 0.04(17%) |
| PCV20 | 0.11(-33%) | 0.09(-27%) | 0.05(-45%) | 0.03(-56%) | 0.04(22%) |
| level 4 MC |  |  |  |  |  |
| No Vaccine | 0.16 | 0.13 | 0.10 | 0.09 | 0.05 |
| PCV7 | 0.11(-33%) | 0.13(0.34%) | 0.10(0.47%) | 0.09(0.61%) | 0.05(1%) |
| PCV13 | 0.11(-32%) | 0.09(-27%) | 0.11(0.77%) | 0.09(1%) | 0.05(2%) |
| PCV15 | 0.11(-32%) | 0.09(-27%) | 0.06(-43%) | 0.09(1%) | 0.05(3%) |
| PCV20 | 0.11(-32%) | 0.09(-27%) | 0.06(-43%) | 0.04(-51%) | 0.05(4%) |
| level 11 MC |  |  |  |  |  |
| No Vaccine | 0.17 | 0.13 | 0.11 | 0.09 | 0.05 |
| PCV7 | 0.11(-32%) | 0.13(0%) | 0.11(0%) | 0.09(0%) | 0.05(0%) |
| PCV13 | 0.11(-32%) | 0.10(-27%) | 0.11(0%) | 0.09(0%) | 0.05(0%) |
| PCV15 | 0.11(-32%) | 0.10(-27%) | 0.06(-42%) | 0.09(0%) | 0.05(0%) |
| PCV20 | 0.11(-32%) | 0.10(-27%) | 0.06(-42%) | 0.04(-51%) | 0.05(0%) |

In a scenario with level 3 MC, akin to the analysis with level 2 MC, we again observe a decrease in the prevalence of *ST*_1_ with the introduction of PCV7. However, in contrast to the previous case, the prevalence of *ST*_2_ rises from 0.119 to 0.1217, amounting to an increase of a meagre 2.256%. Additionally, the prevalence of serotypes *ST*_3_ through *ST*_5_ also increases modestly upon the use of PCV7. Similar to the previous scenario, each vaccine causes an increase in the prevalence of any serotype not included in that specific vaccine.

Increasing to level 4 MC in the model, we observe a similar decrease in the prevalence of *ST*_1_ due to the introduction of PCV7, just like in the previous cases. However, unlike the earlier scenarios, the prevalence of *ST*_2_ actually rises from 0.1299 to 0.1301, representing an increase of a modest 0.1357%. Additionally, the prevalence of serotypes *ST*_3_ through *ST*_5_ also increases even more modestly when PCV7 is administered. Each vaccine leads to a corresponding rise in the prevalence of any serotype not included in that specific vaccine.

This trend motivates the exploration of ST replacement with maximum possible MC in the model. Therefore, we finally examine the scenario in which the model (1) accounts for MC with all 11 serotypes. Upon the introduction of any PCV in this case, each NVT for the respective vaccine continues to exist at the steady-state prevalence observed without vaccination. Prevalence of *ST*_1_ decreases from 0.1667 to 0.1131. PCV13, which has an efficacy equivalent to that of PCV7 (60%), similarly decreases the prevalence of *ST*_1_ to the same value of 0.1131. The effects of PCV15 and PCV20 are similarly illustrated.

A clear pattern of reduced replacement emerges when transitioning from level 2 to level 11 MC. Conversely, the prevalence of NVTs significantly increases as restrictions on co-colonizing serotypes become more stringent.

Notably, the serotypes *ST*_5_, …, *ST*_11_ are classified as NVTs for all four PCVs considered. Therefore, it is beneficial to consolidate these serotypes into a single ST, allowing for the use of a 5-ST model. This approach facilitates an examination of the effects of a reduced number of serotypes on prevalence in the corresponding MC levels.

### 5.1 5-ST model with varying levels of MC

Consolidating *ST*_5_ through *ST*_11_ into a single serotype, denoted as 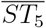, we use the notation 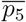 to accordingly denote the sum 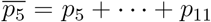. Upon the introduction of PCV15 in the resulting 5-ST model, Figure 3 shows prevalence shifts with increasing levels of MC. Since the properties of *ST*_5_,, *ST*_11_ are identical, *p*_5_ = *· · ·* = *p*_11_. As expected, the increase in prevalence of non-PCV15 serotypes diminishes with increased level of MC. Additionally, a critical trend emerges from our findings: the consolidated serotypes represented by *ST*_5_ contribute to serotype replacement relatively insignificantly compared to other non-PCV15 types. This suggests that the serotypes in the consolidated group maintain a stable presence in the population, thereby isolating the dynamics of replacement to the other non-PCV15 types. It is important to underscore that the total replacement observed within the 5-ST model is lower when compared to the model that includes all 11 serotypes (Table 3). This reduction in replacement implies that modeling with fewer serotypes may mitigate ST replacement, especially when the model can only incorporate low-level MC.

**Table 3:**
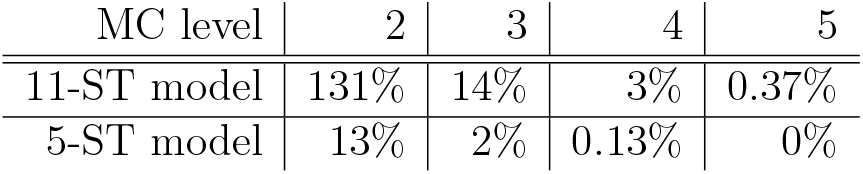
Increase in non-PCV15 ST prevalence with PCV15 in 11-ST and 5-ST models. MC: multi-colonization.

**Figure 3:**
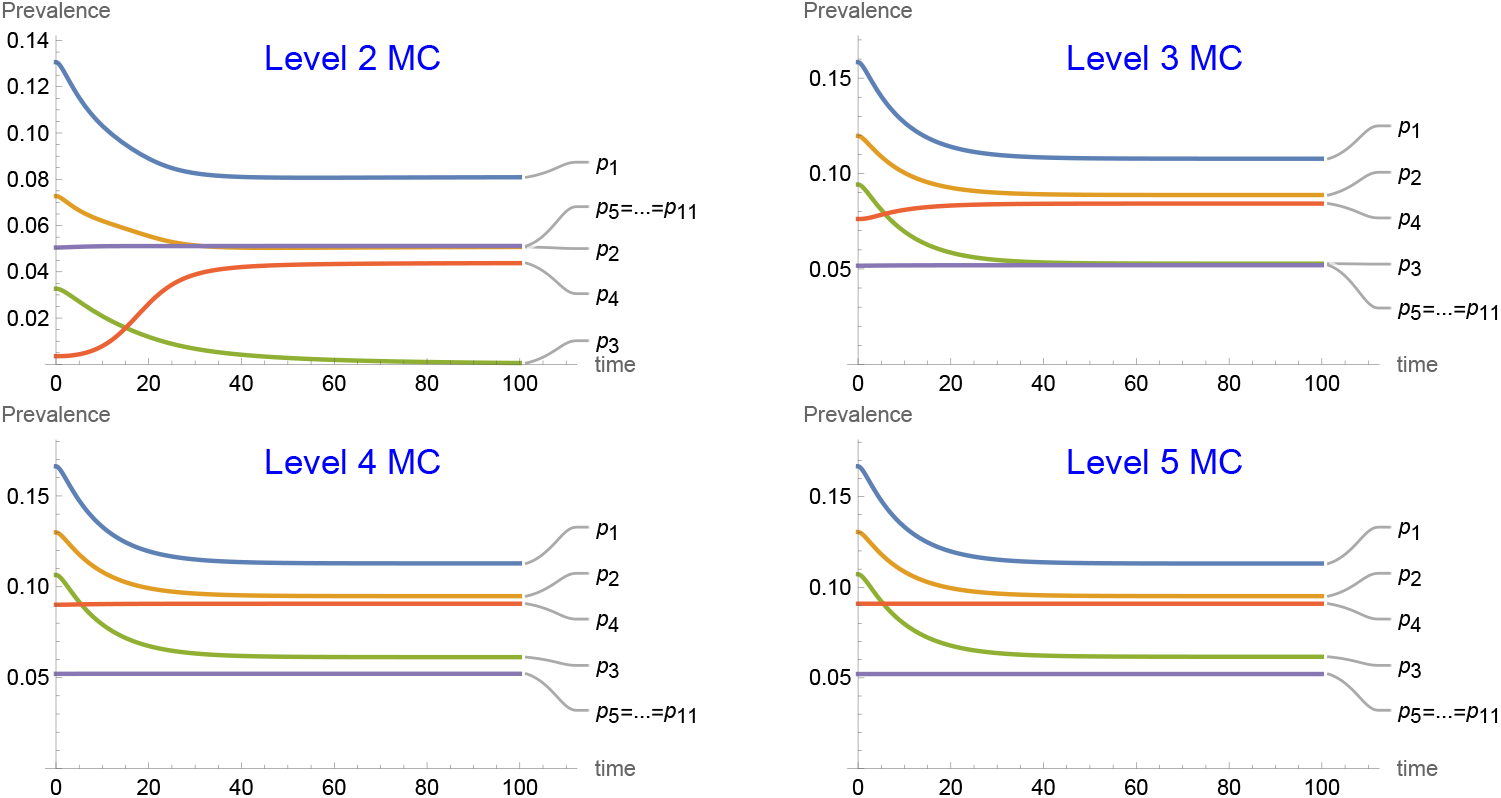
Evolution of serotype prevalence under PCV15 in a 5-ST model by aggregating *ST*_5_, *· · ·, ST*_11_ into *ST*_5_. As a result, *ℛ*_5_ = 1.57. Other parameter values used are as in Figure 2.

The rise in NVT prevalence upon vaccine introduction, despite the absence of direct competition, challenges traditional views on serotype dynamics. It suggests that the observed replacement may not solely stem from metabolic competition among serotypes. Rather, it can also emerge as an artifact due to the constraints inherent in the modeling framework concerning MC. We further show that increased restrictions on the number of co-colonizing serotypes in a model lead to a greater observed rate of replacement.

## 6 Discussion

In this study, we have examined the dynamics of multi-serotype interactions and established that the phenomenon of serotype replacement, often attributed to direct competition among serotypes, can actually emerge from the constraints imposed by modeling assumptions concerning multi-colonization (MC). Our findings elucidate that even in the absence of a priori competition between serotypes, introducing limitations in a model on the number of serotypes that can co-colonize can create conditions conducive to outcomes showing serotype replacement. Consequently, we argue that the observed replacement is not merely an epidemiological phenomenon, but rather an artifact of modeling framework.

The findings from the two-serotype and three-serotype models elucidate important dynamics in serotype interaction under varying assumptions of MC and competition among pneumococcal serotypes. Our results demonstrate that competition between serotypes invariably leads to a reduction in equilibrium prevalence compared to scenarios devoid of any competitive interactions, where the prevalence of each serotype is solely determined by its intrinsic parameters.

In the two-serotype model, the independence in transmission and clearance allows us to infer that the interaction between serotypes is fundamentally governed by the competition parameters. When competition is absent (i.e., with parameters *θ*_1,2_ = *θ*_2,1_ = 1), the prevalence of each serotype remains unaffected by the presence of the other. In contrast, introducing competition diminishes the prevalence of both serotypes, underscoring the significance of competitive interactions in shaping serotype dynamics. The insights from the three-serotype model further clarify the complexities that arise when multiple serotypes coexist. In the absence of metabolic-type competition, we observed that the inclusion of triple colonization in the model led to no possibility of serotype replacement. However, interestingly, even without competition, the prevalence of serotypes at equilibrium is influenced by the presence of other serotypes when MC is limited to double carriage. This finding suggests that, in a three-serotype framework with restricted MC, serotype replacement can still occur, highlighting the nuanced interplay between serotype prevalence and the constraints imposed by MC assumptions in the model.

Our analysis also illustrates a critical aspect of pneumococcal epidemiology: the increase in non-vaccine serotypes as the prevalence of vaccine-targeted serotypes decreases. This phenomenon occurs even in scenarios where direct competition between serotypes is absent (*θ*_*i,j*_ = 1 for all *i, j*). Such dynamics may be attributed to the limitations imposed by the model on the number of serotypes permitted to co-colonize, which accentuates the emergence of non-vaccine serotypes.

The evolution of pneumococcal conjugate vaccines (PCVs), notably the sequential introduction of PCV7, PCV13, PCV15, and PCV20, has transformed the landscape of serotype prevalence. As our results illustrate, the implementation of these vaccines directly correlates with a decline in the prevalence of targeted serotypes. However, in the context of reduced MC, our findings indicate a marked increase in the prevalence of non-vaccine serotypes. For example, when restricting MC to two serotypes, the observed prevalence of non-vaccine types surged significantly. This emergent trend destabilizes traditional perspectives regarding serotype dynamics, implying that the observed serotype replacement may not solely arise from metabolic competition but also from the structural constraints inherent in the modeling framework concerning MC.

The analysis of varying levels of MC underscores a pivotal point: as the number of co-colonizing serotypes in a model decreases, the rate of projected serotype replacement appears to increase. On the other hand, modeling fewer overall serotypes by consolidating non-vaccine types leads to a mitigation of serotype replacement introduced due to MC limitation in the model. These observations point to a critical need for evaluating serotype dynamics in light of MC limitations permitted in modeling studies, thereby urging a deeper investigation into the implications of vaccination strategies on non-vaccine serotype prevalence.

## Data Availability

All data produced in the present work are contained in the manuscript

## 7 Appendix

### 7.1 Mathematical analysis of two-serotype model

#### 7.1.1 Jacobian matrix at the disease-free equilibrium

The Jacobian matrix evaluated at the disease equilibrium is given by:

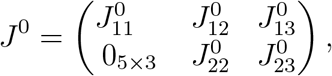

where

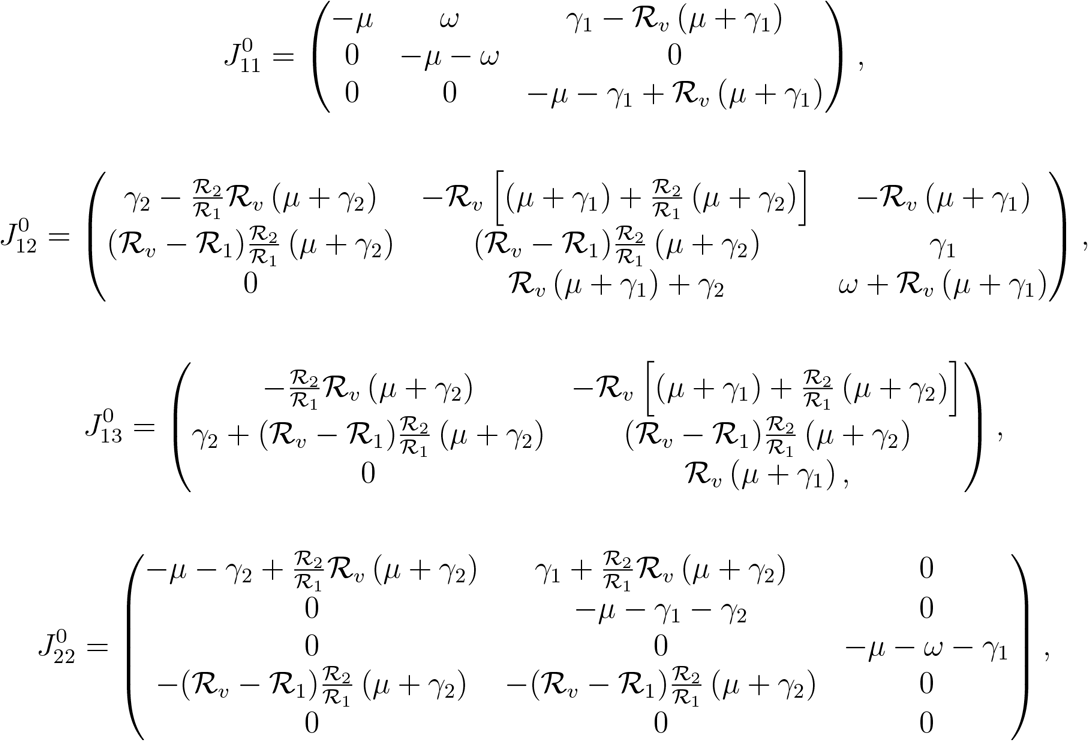

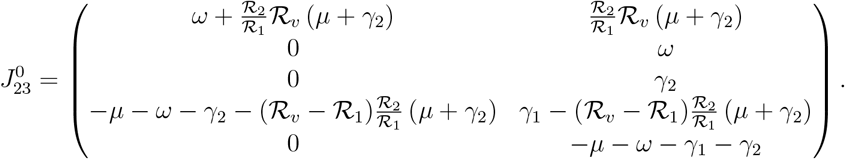

#### 7.1.2 Jacobian matrix at Serotype-1 only boundary equilibrium

The equilibrium where only serotype 1 is endemic exists only if *ℛ*_1_ *>* 1. The Jacobian Matrix evaluated at the at endemic equilibrium of serotype 1 exclusively is given by

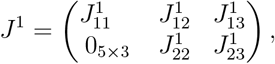

where

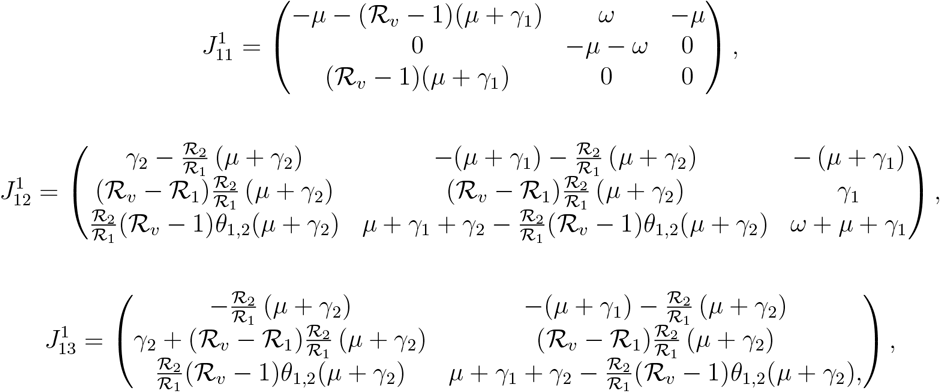

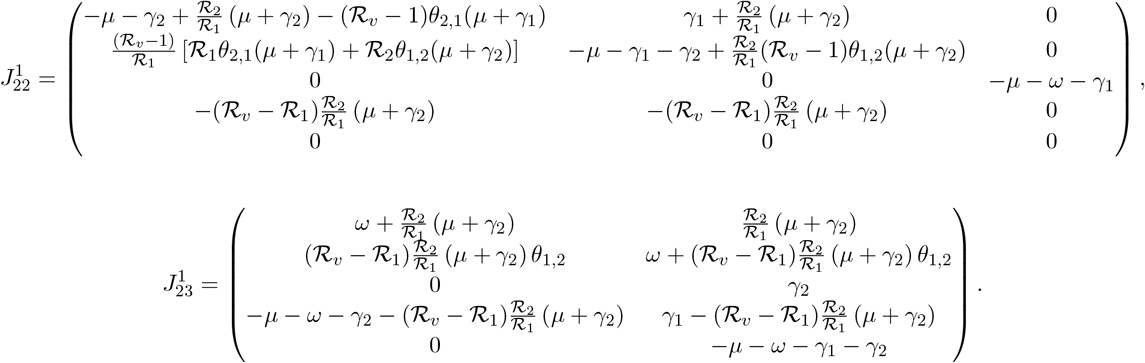

#### 7.1.3 Jacobian matrix at Serotype-2 only endemic equilibrium

The equilibrium where only serotype 2 is endemic exists only if *ℛ*_2_ *>* 1. The Jacobian Matrix evaluated at the at endemic equilibrium of serotype 2 exclusively is given by

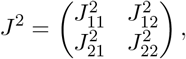

where

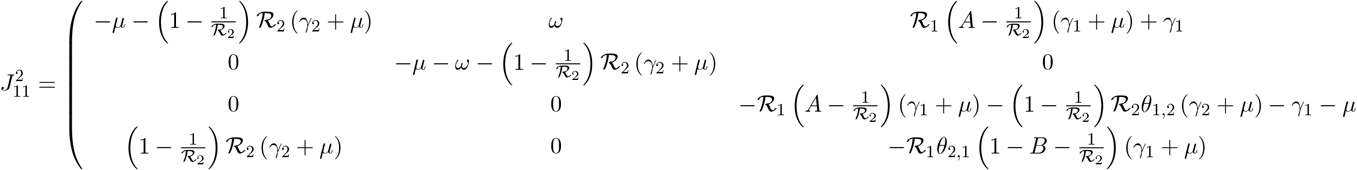

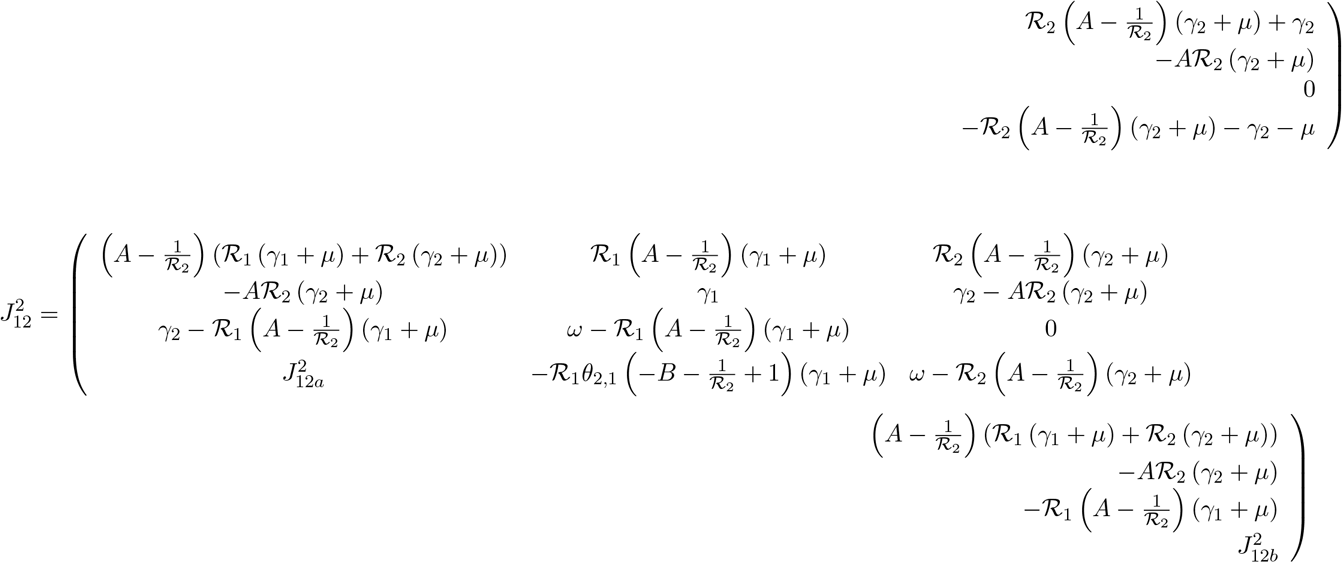

where

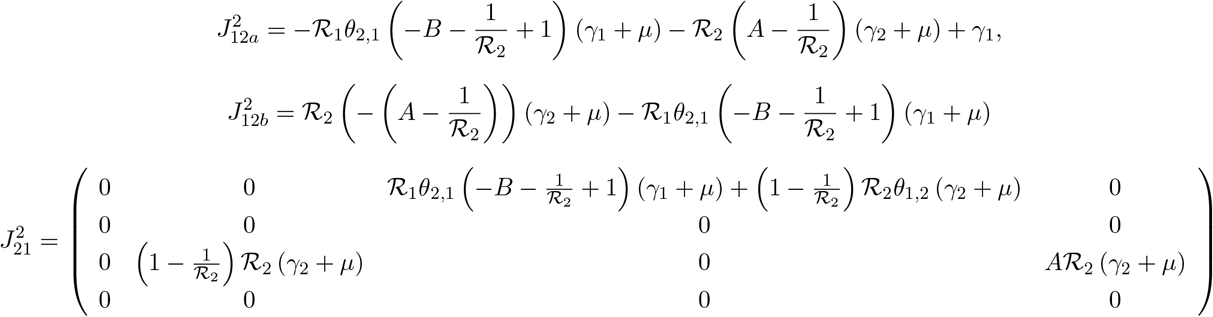

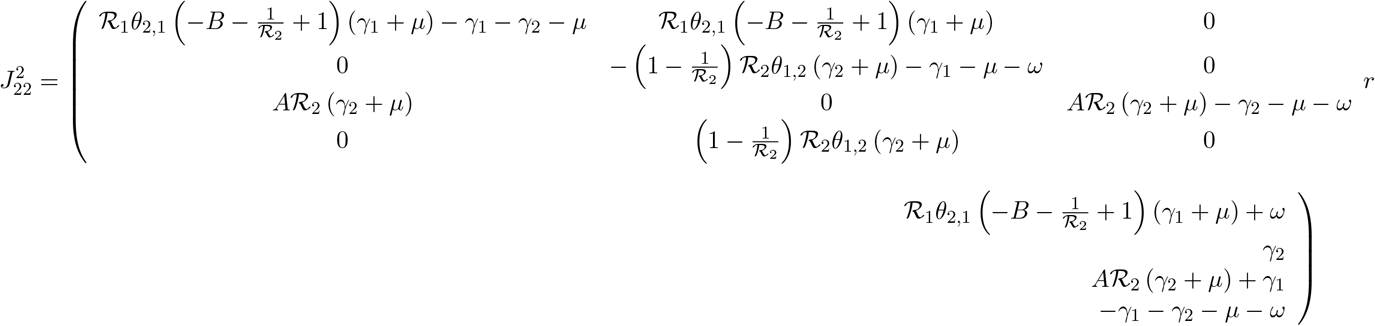

with

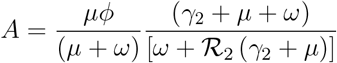

and

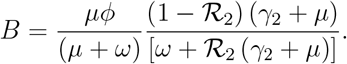

